# Ukrainian refugee integration and flows analysis with an approach of Big Data: Social media insights

**DOI:** 10.1101/2022.04.18.22273958

**Authors:** Tado Jurić

## Abstract

**Background:** The Ukrainian refugee crisis shows a lack of reliable data about refugee flows, demographic structure, and integration. But those data are necessary for the UNHCR and governments in preparing high-quality projections for emergencies and the conditions for the integration of refugees who intend to stay in immigration societies. Although Facebook, Instagram and YouTube are social platforms with the most users, very few studies have been written about their potential for migration studies and integration.

**Objective:** The objective was to test the usefulness of Big Data insights from social network platforms to gain first demographic insights into refugees’ age and gender structure, migration flows, and integration trends.

**Methods:** The primary methodological concept of our approach is to monitor the so-called “digital trace” of refugees left on social networks Facebook, Instagram and YouTube and their geo-locations. We focus on users that use social network platforms in Ukrainian and Russian language. We sampled the data before and during the war outbreak, standardised the data and compared it with the first official data from UNHCR and national governments. We selected specific keywords, i.e. migration and integration-related queries, using YouTube insight tools. Using FB and Instagram, we collected our own data archive because Meta offers data only for the present day with the ability to compare this day with the average of the past 12 months. In the next step, we collected signals that indicate integration willingness.

**Results:** Our approach shows that the number of Facebook and Instagram users is growing rapidly in Ukrainian neighbouring countries and Germany after the war outbreak in Ukraine. Testing performed matches the trend of immigration of Ukrainian refugees in Poland and Germany, as well in the cities and the German *Bundesländer*. The tested correlation between the number of Ukrainian refugees in Poland and FB and Instagram users in Ukrainian in Poland shows that the increase in frequency index is correlated with stepped-up emigration from Ukraine. R2 is 0.1324 and shows a positive correlation, and a p-value is statistically significant. The analysis of the FB group of Ukrainian in the EU shows that those groups can be a valuable source for studying integration. Ukrainians are increasingly expressing interest in learning the German language, which is a good indication of integration willingness. One of the contributions of the second used method, YouTube insights, is that it shows that by searching for video material on the YouTube platform, the intention of users to migrate, or in this case, to flee from Ukraine, can be estimated.

**Conclusion:** The usefulness and the main advantage of this approach are enabling first insights into integration willingness and identification of trends in the movement and intentions of refugees when there is no official data. On the one side, this method allows governments to estimate how many refugees intend to enter the labour market and integrate into the immigration society and, on the other, better respond to the recent humanitarian crisis. Despite its beneficiaries, this approach has many limitations, and there is a need for many more studies to perfect this method.

## Introduction

This study presents a descriptive analysis of Big Data sources, which could help determine refugee flows from Ukraine and give first demographic insights into refugees’ age and gender structure. The second goal is to show that this approach can be useful for assessing the degree of integration of Ukrainian refugees using insights from social media platforms such as Facebook, Instagram and YouTube.

As shown many times over the recent past^1^, the Ukrainian refugee crisis again shows that reliable data about refugee flows and later about their integration would have assisted UNHCR and governments in preparing high-quality projections for emergencies and the conditions for the integration of those who intend to stay in immigration societies. ^2^ However, such data are often unavailable or only available with a considerable time lag.^3^

Although Facebook (FB), Instagram and YouTube are the most used social platform^4^, very few studies have been written about their potential for migration studies and integration insights.^5^ We will show that there are several approaches that insights obtained from digital traces left on social networks can be used to identify and model migratory movements and integration of immigrants, in this case, refugees. Because refugees are a vulnerable group and social research with a typical approach of interviews and surveys at this time is not feasible and, in some cases, inappropriate due to the traumatisation of respondents – this is another advantage of this method.

Meta Business Suite can provide insights into geo-location and particular interests of the observed population based on many signals such as likes, pages visited and specific cultural interests. We will show that these insights can be used to modelling migration flows and illuminate the cultural assimilation and integration of a particular group of emigrants into other societies.^6^ The same approach can be used to analyse digital tracks on Instagram, while insights from YouTube, the second most visited social network, can be obtained by analysing keyword searches.

We will compare the data obtained from the analysis of social networks with official UNHCR data and available data from EU governments, test the use-value of Big Data about refugees’ integration willingness, and model further trends related to the refugee crisis in Ukraine.

The structure of the paper is as follows: after briefly showing the results of previous studies, we explain the methodology of gaining insights from social networks platforms for migration and integration research and will show the limitations of this method. In the section on results, we reveal the results achieved via this approach and discuss how to estimate refugee migration flows with social media networks and what insights about refugees’ integration willingness can we gain from social media.

## I. Facebook, Instagram, and YouTube insights as a method for determining refugee migration flows

Traditional data sources generally fail to provide statistical information on migration and refugee flows quickly^7^, which is evident during the current refugee crisis. A study by Singh et al. (2019) on internally displaced person’s movements between provinces in Iraq showed that a mix of social media data and traditional register data improves the predictive quality compared to predictions based on register data alone.^8^ Several studies have used Big Data sources to analyse migration, i.e. refugee flows.^9^ It is proven that the refugees are more interested in information from the Internet than the average and that they generally use smartphones during migration.^10^ Therefore, Big Data has the potential for governments and UNHCR to generate better statistics and improve their early warning systems. ^11^

Previous research has proven that Big Data can be used to study migration, but there are still significant methodological issues and scepticism regarding the feasibility of using alternative data sources.^12^ Zagheni (2020)^13^ was the first to show that FB can be an invaluable source of data unused for demographic research and that it offers a set of data that is a kind of “constantly updated census”^14^, but many authors also questioned this approach.^15^ For a detailed review of the literature and the current discussion on the application of Big Data in migration and refuge studies, see our previous study (Jurić, 2022) *Predicting refugee flows from Ukraine with an approach to Big (Crisis) Data*.^16^

Facebook, Instagram, and YouTube can also serve as reliable tools to monitor the degree of integration of refugees into incoming society, primarily through the analysis of interests.^17^ For example, Dubois et al. (2018) estimated the level of assimilation of Arabic-speaking migrants in Germany based on FB data.^18^ Herdagdelen et al. (2016) used FB to describe the composition of immigrants’ social networks in the United States.^19^ Zagheni et al. (2018) tested this model in the US and Jurić (2022) in Germany.^20^

FB and Instagram (now “Meta”^21^) have devoted significant resources to assessing their users’ demographic characteristics, behaviours, and interests to improve targeted advertising, which is their primary source of revenue. For this purpose, FB has specifically developed a platform called Adverts Manager (FB ADS) and integrated it in 2022 with Instagram to *Meta Bussines Suite*^22^, allowing advertisers to select detailed user characteristics to which their ads should be displayed.^23^ Socio-demographic data includes data such as the geo-location, age or gender of users, networks of friends and related websites, as well as expressed interests. For example, FB and Instagram support the display of advertisements exclusively to Ukrainian emigrants who are adults and are now in Berlin or Munich. Due to such a possibility, Meta tools offer beneficial insights for demographic research.^24^

The primary method used for analysing YouTube is based on search language and geo-location. The insight tools of *Google Analytics* allow analysis according to the country where the specific group uses these applications and the language of use. Jurić (2022) has shown that would-be migrants often use online searching to get answers about the country they plan to emigrate to^25^, and Wanner (2020) that Google is the first source of information for most users planning emigrate.^26^ In our previously mentioned study (Jurić 2022) ^27^, we tested the predictive capacity of Google Analytics regarding refuge flow from Ukraine and proved that these insights give reliable data and can be helpful in modelling future trends. The difference from the previous approach in this paper is that we focus on the Google social network YouTube and not the Google search engine as well, as mentioned, FB and Instagram.

In previous studies^28^, there were important limitations that each of these searches was conducted for its reason and did not answer researchers’ questions, so searching the term “Poland” or watching a video about Poland cities was not necessarily an implication that someone wanted to move to Poland. However, in this refugee crisis, we can start from a firmer assumption that informing by watching videos on YouTube Platform from Ukraine about Poland or German cities, cultures and way of life gives more reliable indications of the intention to migrate, i.e. flee to these countries. Compared to data from Meta, the advantage of YouTube is that limitations related to penetration rates and double fake accounts are not prevalent.^29^

## II. SOCIAL MEDIA AS A METHOD FOR DETERMINING AND MEASURING REFUGEE INTEGRATION TRENDS

Social integration is the process during which newcomers are incorporated into the social structure of the host society. ^30^ This process is a dynamic and multifaceted two-way process that requires efforts by all parties concerned, and there is no “one-size-fits-all” approach to integration. The situation of refugees must be analysed in the context of the respective host society and concerning the living and working conditions of nationals.^31^ Strang and Ager show that prevailing notions and understanding of nationhood and citizenship determine understandings of integration, and argue that this powerfully shapes the social space available to refugees regarding belonging.^32^ According to UNHCR, it is in the best interests of both the host society and refugees to promote a reception policy with a long term perspective.^33^

Jurić (2022) has proven that through the analysis of FB interests, the degree of integration of emigrants into incoming society can be monitored quite reliably.^34^ The basic idea is that if the interests were similar, one could speak of successful integration or cultural assimilation.^35^ For example, the interests of Ukrainian refugees in Germany unquestionably indicate that learning the German language is extremely important to them, which is an excellent insight into the integration will of this group (see Results). By measuring integration willingness with these tools, we have paid attention to indicators that include education, employment and language, as these indicators show critical integration fields.

The advantage of FB and Instagram over YouTube is that they provide more precise socio-demographic data and insights^36^ into the numerous interests of Ukrainian emigrants, which can be used to monitor their integration. On the other side, the advantage of YouTube is that it better perceives the user’s intention. If sufficiently predictive terms are analysed using YouTube, numerous intentions of the observed population can be revealed (more details in the next section).

Regarding the general penetration of social media platforms among the Ukrainian population used in this analysis, it can be seen that the first four social media are Facebook, which is using 57.6%, YouTube, 12.3%, Twitter, 8.69%, and Instagram, 8.04%^37^ The most used is FB which in February 2022 counted over 26 million Facebook users in Ukraine.^38^ Compared to the EU average^39^, Ukrainians are using FB more and YouTube less.

## III. METHODOLOGY

The primary method we used for analysing YouTube is based on search language and geo-location, and for Facebook and Instagram, on geo-location and the primary language of users. Both tools allow analysis according to the country where the specific group uses these applications. The control mechanism for testing this sort of data was performed by comparing those insights with the official databases from UNHCR and national governments.

Using YouTube, one should especially take care to choose specific keywords, i.e. migration and integration-related queries. According to Wanner (2020), Wilde et al. (2020) and Böhme et al. (2020)^40^, one can expect to find a relationship between the intention of the users and particular behaviours. Therefore, we have focused on monitoring the terms that indicate the intention of refugees from Ukraine to move to Germany. For this purpose, we followed the expressed interests related to life in Germany, videos about German cities, video lessons that offer the opportunity to learn German, etc. (see Results). However, the YouTube search Index insights do not show the exact number of searches in a specific country, so with this tool, the exact number of potential emigrants, i.e. refugees, cannot be calculated, but the increase of the trend can be noticed very precisely.^41^

The basic procedure used so far to estimate how *Meta* recognises migrants was to monitor individuals who qualify as “expats” on their FB profiles.^42^ However, this method cannot be used in the case of Ukrainian refugees because, in a situation of trauma to which the refugee is exposed, few people think about changing the entered data on their FB profile. To track FB and Instagram users by Ukrainian language and geo-location, we have collected data about users in the countries of Ukrainian neighbourhood and Germany every day since the outbreak of the war in Ukraine. This procedure was necessary because *Meta* offers data only for the present day with the ability to compare this day with the average of the past 12 months. Therefore, it is important to note that the researcher has to keep their own data archive – which is a significant limitation compared to data from the YouTube platform, which provides insights into every date from 2008 to date. In the next step, we collected signals indicating the willingness to learn German and job search as key determinants indicating integration willingness.

The method for estimating integration willingness evolved as follows: we collected the state-level estimates of Ukrainian refugees from the *Meta* Business Suite database and FB Marketing API^43^. Since *Meta* can also provide insights into specific interests of the observed population, based on likes, pages visited, and other signals^44^, we used this instrument to analyse their integration willingness through observation of signs that indicate language learning of incoming societies and searching for a job. Of course, it is too early to get deeper insights because refugees are in new communities only for a short time (see section Restrictions).

We collected the data for the ten most shown interests of refugees in Germany and compared this data with data from the German population. As already mentioned, if the interests were similar, one could speak of successful integration. Of course, this assumption cannot be confirmed in such a short time, but it will be useful in future studies. An additional mechanism that can be used in the future is to check the reviews that FB and Instagram users left when reporting their geo-locations, like visited events, etc. It is to note that we use exclusively aggregate data from which the identity of an individual user cannot be reconstructed.

By using this method, it is most important to pay attention to the limitations that can lead to misinterpretations of the results. *Meta* states that estimates are not intended to be consistent with population, census, or other sources. These estimates depend on how many accounts an individual has on the FB and Instagram Apps and Services; how many transient visitors are in a specific geographic location, and demographic information provided by FB users themselves.^45^ A serious limitation of our entire approach is that many Ukrainians use these social platforms in other languages, such as Russian and English (see section Limitations).

## IV. REFUGEE FROM UKRAINE IN THE EU

The Ukraine war started on 24 Feb 2022. ^46^ According to data from UNHCR, the speed of the exodus is already more extensive than the migration crisis of 2015, when 1.3 million asylum seekers entered Europe.^47^ Our previous study showed that there would be at least 5.4 million refugees.^48^

The IOM estimates that more than half of the internally displaced people are women,^49^ and according to UNICEF, more than half of all children in Ukraine have been forced to leave their homes.^50^ More than 60% of households are travelling with children, and of the nearly 10 million people displaced within and outside Ukraine, 186,000 are nationals of a third country.^51^ Almost 30% of internally displaced had come from Kyiv, more than 36% had fled from the east of Ukraine; 20% had come from the north, and nearly 40% were now in the west of Ukraine. The European Commission adopted a right to stay approach, allowing Ukrainian refugees to travel freely within the EU’s borders, remain where they choose, work legally, and access social services.^52^

Most Ukrainians fled to Poland, which welcomed 2,405,703 people (01. April), followed by Romania, with 623,627.^53^ The number of refugees is also high in other neighbouring countries, Hungary, Slovakia and Moldova. Of the other EU countries, most refugees went to Germany, 310,000.^54^

As time goes on, more and more refugees are coming to other EU countries. It is also noticeable that more and more refugees decide not to stay in the countries they first arrive in from Ukraine. For example, of 140,000 people who came to Romania during the first eight days of the war, 60,000 travelled to other countries.^55^

However, it is complicated to say precisely how Ukrainian refugees are distributed in individual countries, as there is no exact data than just estimates. Studies were only being carried out in Germany at this time.^56^ Our previously mentioned study predicted that one-third of all refugees would move to Germany in 2022.^57^

A Survey of Ukrainian war refugees done by the German Federal Ministry of the Interior from (n = 1,936) showed that 42% of those questioned want to stay in Germany and not return to Ukraine, 32% expect to return to Ukraine soon, and 25% have not yet a plan.^59^

## V. RESULTS AND DISCUSSION

### V.1. Social media as a source of migration data

Before we look at the results obtained on social networks, we briefly present the official UNHCR data to compare these two data sources. UNHCR states that on 1 April 2022, there was 2,405,703 Ukrainian refugees in Poland, 379,988 in Hungary, 292,309 in Slovakia, 390,187 in Moldova, 623,627 in Romania, and 737,000 in other Western European countries. Of the other not directly neighbouring countries, the most, 310,000, arrived in Germany.^60^

Our approach with social media platforms shows that the number of FB and Instagram users is growing rapidly in Poland, Slovakia, Hungary, Moldova, Romania, and Germany after the war outbreak in Ukraine.

From Table 3, we can conclude that in this first phase, FB and Instagram, on average, register 17% - 20% of the refugee population in these countries. Below (Figure 2) are two examples of the apparent increase in the number of FB and Instagram platforms users, which coincide with the trend of immigration of Ukrainian citizens in Poland and Germany due to forced migration.

**Table 1.** Refugees from Ukraine in neighbouring countries and West European countries (UNHCR, 01. April 2022)

| Datum | Poland | Hungary | Slovakia | Moldova | Romania | Other European c. (West) |
| --- | --- | --- | --- | --- | --- | --- |
| 28. Feb 2022 | 280.000 | 85.000 | no exact data available | no exact data available | no exact data available | 34,600 |
| 04. Mar 2022 | 756,303 | 157,004 | no exact data available | no exact data available | no exact data available | 133,876 |
| 08. Mar 2022 | 1,204,403 | 191,348 | no exact data available | no exact data available | no exact data available | 210,239 |
| 11. Mar 2022 | 1,524,903 | 225,046 | no exact data available | no exact data available | no exact data available | 282,497 |
| 01. April 2022 | 2,405,703 | 379,988 | 292,309 | 390,187 | 623,627 | 737,000 |
| **Total: 4,137,842 (Data as 01 Apr 2022)** |  |  |  |  |  |  |

<sup>53</sup> UNHCR, https://data2.unhcr.org/en/situations/ukraine
<sup>54</sup> Medendienst Integration, https://mediendienst-integration.de/migration/flucht-asyl/ukrainische-fluechtlinge.html (01.04.2022)
Author's construction based on UNHCR data. Source: UNHCR, https://data2.unhcr.org/en/situations/ukraine and UNHCR, Ukraine Situation Regional Refugee Response Plan, https://data2.unhcr.org/en/situations/Ukraine.
Note: About 400.000 refugees fled to Belarus and Russia.

**Table 2.** Other EU countries received more than 10,000 Ukrainian refugees (the government reported figures on 01.04.2022)

| Country | Number | Portugal | 25,000 |
| --- | --- | --- | --- |
| Czechia | 300,000 | Denmark | 24,000 |
| Germany | 310,000<sup>58</sup> | Sweden | 21,262 |
| Bulgaria | 136,412 | Netherlands | 21,000 |
| Italy | 79,047 | Croatia | 15,000 |
| France | 45,000 | Finland | 15,000 |
| Lithuania | 38,600 | Latvia | 12,000 |
| Estonia | 25,720 | Norway | 11,000 |
| Spain | 25,000 | Austria | 150.000 |

Source: https://data2.unhcr.org/en/situations/Ukraine and the Government data: 1) Council of Ministers of the Republic of Bulgaria (29 March 2022). "България с грижа за хората от Украйна", 2.) Italian Government (2 April 2022). "Profughi dall'Ucraina: 80.622 le persone arrivate finora in Italia", 3.) IOM, https://www.iom.int/news/almost-65-million-people-internally-displaced-ukraine-iom 4.) Vlada.hr, https://vlada.gov.hr/vijesti/za-ukrajinske-izbjeglice-osigurano-najvise-3600-kuna-mjesecno-za-smjestaj-i-rezije/35122

**Table 3.** The number of Facebook and Instagram users in Ukrainian (Ukrainian refugees) in Poland, Slovakia, Hungary, Moldova, Romania and Germany (07. March – 10. April 2022)

| Year 2022 | Poland | Slovakia | Hungary | Moldova | Romania | Germany |
| --- | --- | --- | --- | --- | --- | --- |
| 07. March | 282.000 | no data available | no data available | no data available | no data available | 51.400 |
| 12. March | 324.200 | no data available | no data available | no data available | no data available | 58.600 |
| 13. March | 329.000 | 2.200 | 1.900 | no data available | no data available | 59.700 |
| 23. March | 367.000 | 7.600 | 6.700 | 9.800 | no data available | 66.800 |
| 02. April | 396.500 | 17.800 | 16.400 | 28.800 | 2.600 | 72.700 |
| 10. April | 396.500 | 17.800 | 16.400 | 28.800 | 2.600 | 72.700 |

Source: Meta Business Suite, data collected and systematised by the author.
Note: The maximum estimate value is displayed. Data for Slovakia, Hungary, Moldova and Romania were not reported because, at the time of the study, they did not exceed 1,000 users, which is the minimum for Meta to register.

**Figure 1:**
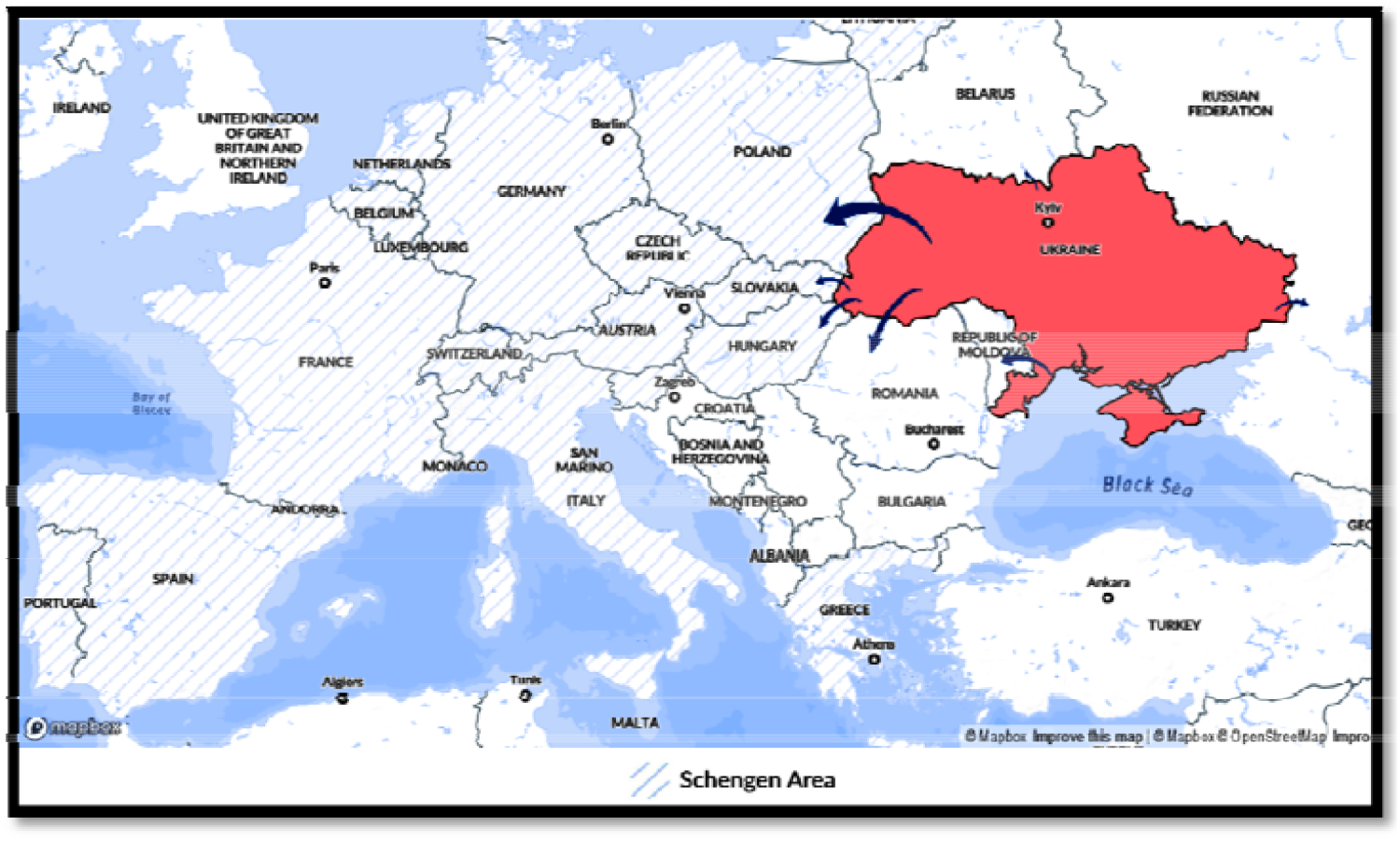
Refugees fleeing Ukraine (24. Feb 2022 – 01. Apr 2022) Source: Microsoft BI, https://app.powerbi.com/view?r=eyJrIjoiNWFlMTQyNjktNzc0ZC00NjZjLWJiYzQtNTZjYzU1ZTIzOTdhIiwidCI6ImU1YzM3OTgxLTY2NjQtNDEzNC04YTBjLTY1NDNkMmFmODBiZSIsImMiOjh9&pageName=ReportSection9165391ce39c58c87c01 (01.04.2022)

**Figure 2.**
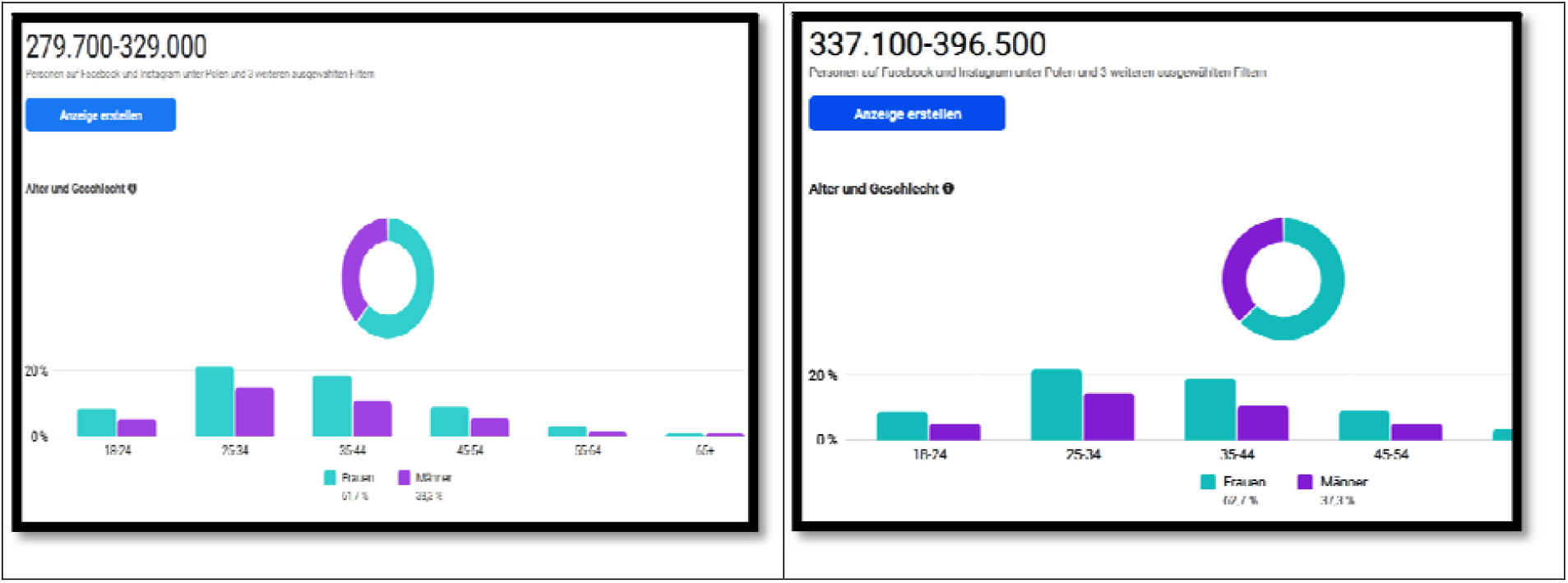
Facebook and Instagram users in Ukrainian (Ukrainian refugees) in Poland, (13. March and 02. April 2022) Source: Meta Business Suite

Figure 2 shows that on 13. March estimated population from Ukraine in Poland (FB and Instagram users in Ukrainian) was 279.700 – 329.000, and 19 days after (on 02. April 2022), the estimated population was 67.500 higher. It is to note that in the first period in which we monitored this data (from 01. March to 05. March), we noticed every day that the number of users of FB and Instagram in Ukrainian in Poland grew by 7,000 to 10,000, while in Germany, it grew by 1,000 to 2,000 new users. Later, this trend was mitigated. Therefore, it is crucial to recheck this data soon (see section Limitations).

Figure 3 shows that the estimated population from Ukraine in Germany (FB and Instagram users in Ukrainian) grew in 19 days from 49.800 – 58.600 to 61.800 – 72.700. These examples unequivocally confirm that FB and Instagram correctly notice trends and that our method can therefore be helpful in migration and refugee studies. Moreover, the control mechanisms we applied testing the model over the same period in terms of migration from Sweden to Germany, Spain to France and Italy to Austria show no rapid increase in value.

**Figure 3.**
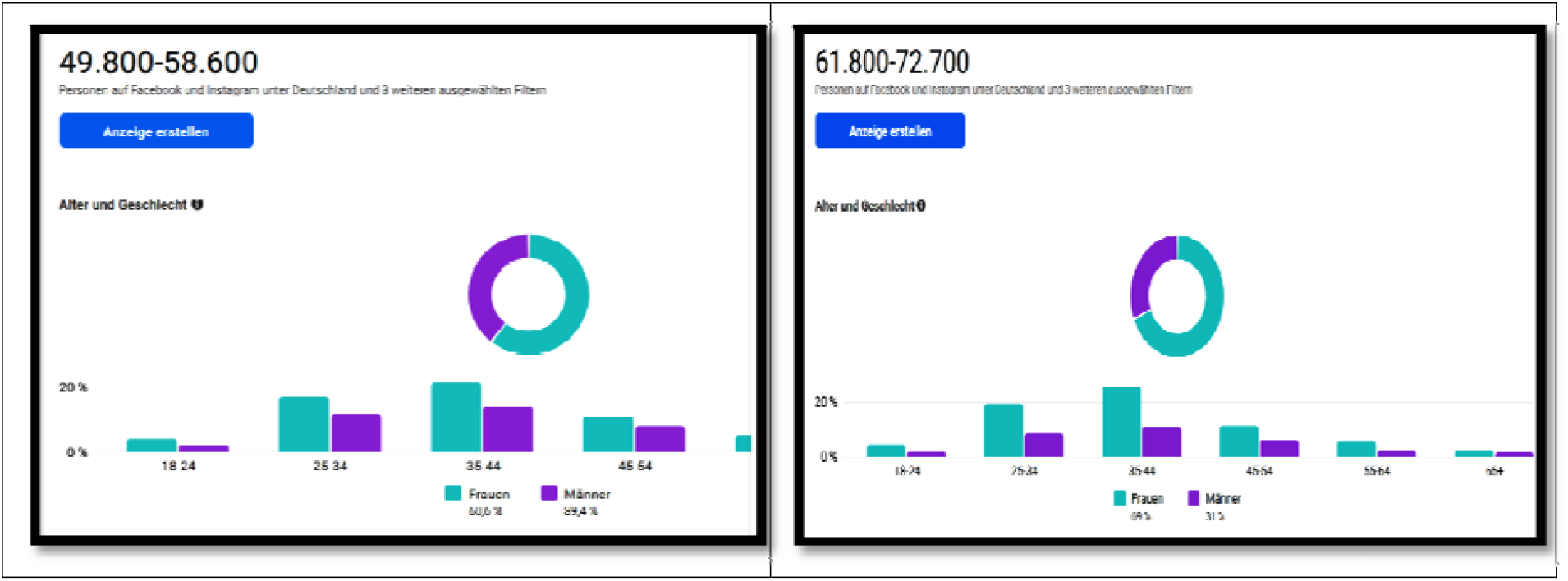
Facebook and Instagram users in Ukrainian (Ukrainian refugees) in Germany, (13. March and 02. April 2022) Source: Meta Business Suite

The study from *Mediendienst Integration*^61^ shows that refugees in Germany mainly arrive in Berlin and Hamburg and are then distribute to other federal states. 42 % are staying in large cities - especially in Berlin (14 %), Munich (5 %) and Hamburg (3 %).^62^ Table 4 shows that data obtained by Facebook and Instagram coincide with the official data from Germany. For other countries, in the absence of official indicators, we have compared the data from our approach with the data reported in the media with the same results.^63^

**Table 4.**
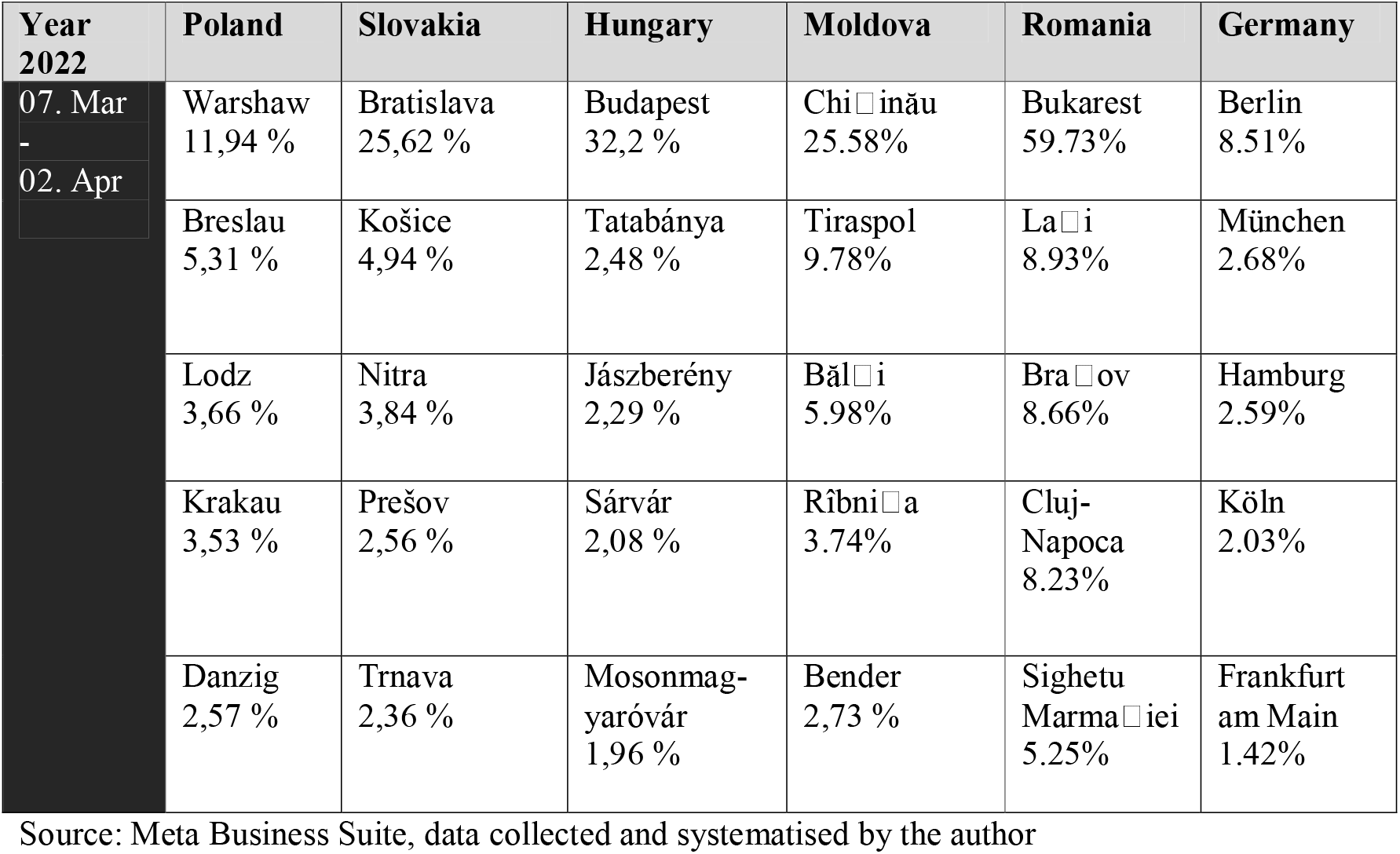
Cities with the most Facebook and Instagram users in Ukrainian (Ukrainian refugees) in Poland, Slovakia, Hungary, Moldova, Romania and Germany.

| Year 2022 | Poland | Slovakia | Hungary | Moldova | Romania | Germany |
| --- | --- | --- | --- | --- | --- | --- |
| 07. Mar - 02. Apr | Warsaw<br>11,94 % | Bratislava<br>25,62 % | Budapest<br>32,2 % | Chişinău<br>25.58% | Bukarest<br>59.73% | Berlin<br>8.51% |
|  | Breslau<br>5,31 % | Košice<br>4,94 % | Tatabánya<br>2,48 % | Tiraspol<br>9.78% | Laşi<br>8.93% | München<br>2.68% |
|  | Lodz<br>3,66 % | Nitra<br>3,84 % | Jászberény<br>2,29 % | Bălţi<br>5.98% | Braşov<br>8.66% | Hamburg<br>2.59% |
|  | Krakau<br>3,53 % | Prešov<br>2,56 % | Sárvár<br>2,08 % | Rîbniţa<br>3.74% | Cluj-Napoca<br>8.23% | Köln<br>2.03% |
|  | Danzig<br>2,57 % | Trnava<br>2,36 % | Mosonmagyaróvár<br>1,96 % | Bender<br>2,73 % | Sighetu Marmaţiei<br>5.25% | Frankfurt am Main<br>1.42% |

Source: Meta Business Suite, data collected and systematised by the author
<sup>61</sup> Mediendienst Integration, https://mediendienst-integration.de/migration/flucht-asyl/ukrainische-fluechtlinge.html (10.04.2022)
<sup>62</sup> Bundesministerium des Innern und für Heimat, Pressemitteilung, 04.04.2022, Befragung ukrainischer Kriegsflüchtlinge.

The German Federal Ministry of the Interior study shows that the refugees primarily went to places where there were friends or relatives or where they had hopes of finding work.^64^ For 82% of those questioned, Germany was the primary escape destination, and the most common escape route was via Poland (65%). The majority of the war refugees from Ukraine questioned were women (84%), of whom 58% fled with their children, and only 17% came alone and unaccompanied (mostly older people). The average age of the interviewed refugees is 38.2 years.^65^ For the most part, these data also coincide with the data obtained through the analysis of social networks. A significant limitation here is that children under the age of 18 cannot be included in the research and that the official data are not sampled for all countries, so it was not possible to carry out a complete comparison (see section Restrictions). However, what our approach unquestionably correctly observes are the recent trends.

Table 5 shows that the users are most represented in the age group 25-34 and 35-44 and that there are almost twice as many women in this age group - which overlaps with the mentioned German study data. According to Meta, the share of women in the refugee population from Ukraine is 62.7% in Poland and 69% in Germany. When we set the filter to limit the specific interests that only Ukrainians in Poland could have and keep the Ukrainian language option, we get a more accurate picture showing a demographic structure with 73.1% of women.

**Table 5.**
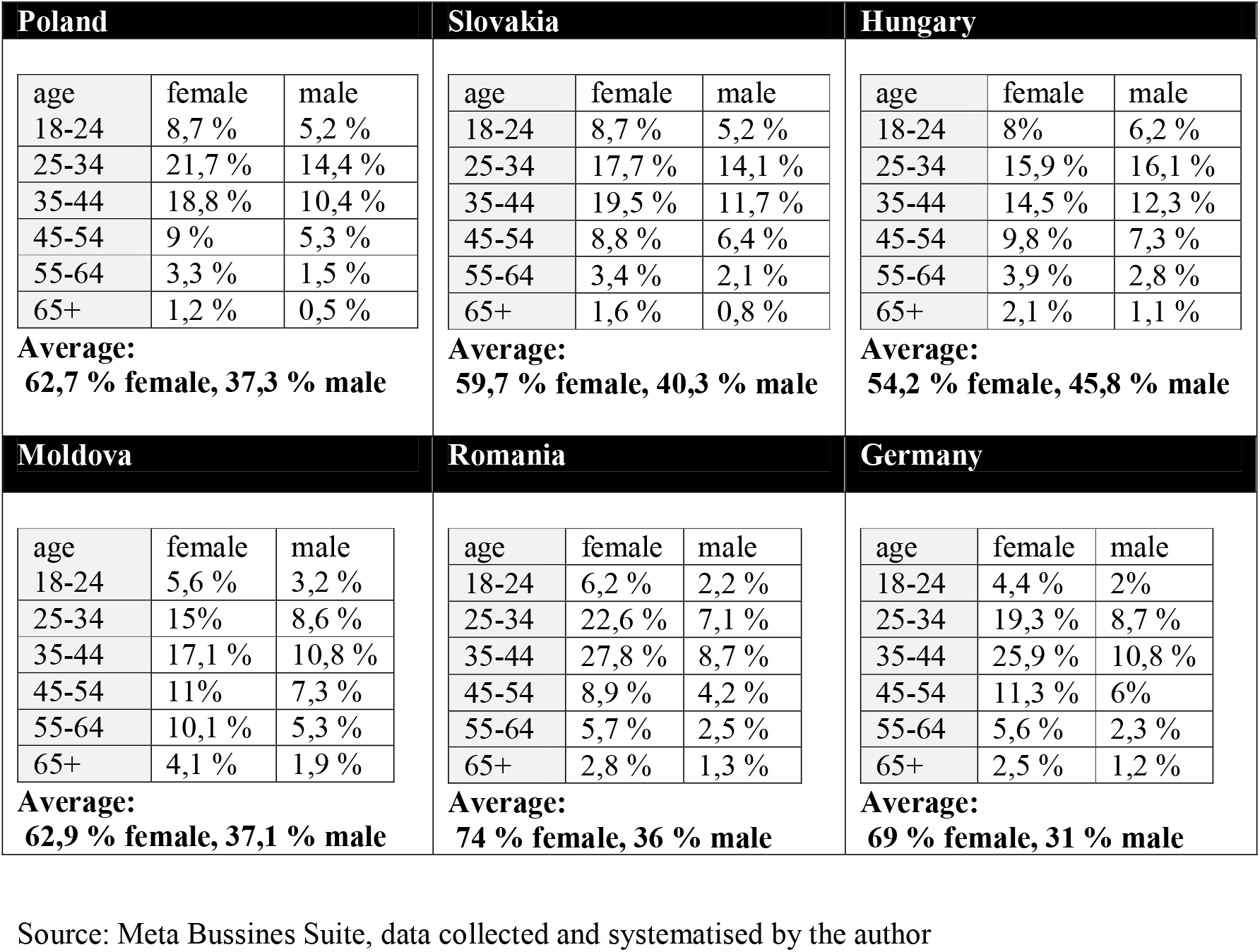
Age and sex of Facebook and Instagram users in Ukrainian (Ukrainian refugees) in Poland, Slovakia, Hungary, Moldova, Romania and Germany (01. Mar -02. April)

| Poland |  |  | Slovakia |  |  | Hungary |  |  |
| --- | --- | --- | --- | --- | --- | --- | --- | --- |
| age | female | male | age | female | male | age | female | male |
| 18-24 | 8,7 % | 5,2 % | 18-24 | 8,7 % | 5,2 % | 18-24 | 8% | 6,2 % |
| 25-34 | 21,7 % | 14,4 % | 25-34 | 17,7 % | 14,1 % | 25-34 | 15,9 % | 16,1 % |
| 35-44 | 18,8 % | 10,4 % | 35-44 | 19,5 % | 11,7 % | 35-44 | 14,5 % | 12,3 % |
| 45-54 | 9 % | 5,3 % | 45-54 | 8,8 % | 6,4 % | 45-54 | 9,8 % | 7,3 % |
| 55-64 | 3,3 % | 1,5 % | 55-64 | 3,4 % | 2,1 % | 55-64 | 3,9 % | 2,8 % |
| 65+ | 1,2 % | 0,5 % | 65+ | 1,6 % | 0,8 % | 65+ | 2,1 % | 1,1 % |
| **Average:**<br>**62,7 % female, 37,3 % male** |  |  | **Average:**<br>**59,7 % female, 40,3 % male** |  |  | **Average:**<br>**54,2 % female, 45,8 % male** |  |  |
| Moldova |  |  | Romania |  |  | Germany |  |  |
| age | female | male | age | female | male | age | female | male |
| 18-24 | 5,6 % | 3,2 % | 18-24 | 6,2 % | 2,2 % | 18-24 | 4,4 % | 2% |
| 25-34 | 15% | 8,6 % | 25-34 | 22,6 % | 7,1 % | 25-34 | 19,3 % | 8,7 % |
| 35-44 | 17,1 % | 10,8 % | 35-44 | 27,8 % | 8,7 % | 35-44 | 25,9 % | 10,8 % |
| 45-54 | 11% | 7,3 % | 45-54 | 8,9 % | 4,2 % | 45-54 | 11,3 % | 6% |
| 55-64 | 10,1 % | 5,3 % | 55-64 | 5,7 % | 2,5 % | 55-64 | 5,6 % | 2,3 % |
| 65+ | 4,1 % | 1,9 % | 65+ | 2,8 % | 1,3 % | 65+ | 2,5 % | 1,2 % |
| **Average:**<br>**62,9 % female, 37,1 % male** |  |  | **Average:**<br>**74 % female, 36 % male** |  |  | **Average:**<br>**69 % female, 31 % male** |  |  |

Source: Meta Bussines Suite, data collected and systematised by the author
<sup>64</sup> Bundesministerium des Innern und für Heimat, Pressemitteilung, 04.04.2022, Befragung ukrainischer Kriegsflüchtlinge, https://www.bmi.bund.de/SharedDocs/downloads/DE/veroeffentlichungen/nachrichten/2022/umfrage-ukraine-fluechtlinge.pdf?__blob=publicationFile&v=2
<sup>65</sup> Bundesministerium des Innern und für Heimat, Pressemitteilung, 04.04.2022, Befragung ukrainischer Kriegsflüchtlinge.

Regarding the cities with the highest activity of Ukrainians in Germany and Poland, we can see that these findings again correspond to what official German statistics are reporting^66^; most Ukrainians are in Berlin, Munich and Hamburg.

As for the change in the number of users, from Table 6, it can be noticed that the index change from 6 April to 15 April occurred only in Berlin (500 new users). According to media data, this data certainly does not correspond to the real situation because the number of newly arrived refugees is much higher.

**Table 6.** Overall assessment of Ukrainian refugees by cities in 2022 according to FB.

| City | 06. April 2022 | 15. April 2022 |
| --- | --- | --- |
| Berlin | 9,300 | 9,800 |
| Muenchen | 3,500 | 3,500 |
| Hamburg | 2,800 | 2,800 |
| Koeln | 2,200 | 2,200 |
| Warschau | 59,000 | 59,000 |
| Breslau | 28,000 | 28,000 |
| Krakau | 19,700 | 19,700 |

**Table 7.** German and Poland cities with the highest activity in the Ukrainian language on FB and Instagram, April 2022.

|  |  |  |  |
| --- | --- | --- | --- |
| Berlin | 8,39 % | Warschau | 11,88 % |
| München | 2,65 % | Breslau | 5,29 % |
| Hamburg | 2,61 % | Krakau | 3,51 % |

Source: FB API, data collected and systematised by the author

We also checked the number of Ukrainian refugees in German federal states. The highest result of using FB and Instagram in Ukrainian is seen in the Bundesland Bavaria, Berlin and Hamburg (12,300 in Bavaria; 9,800 in Berlin and 2,800 in Hamburg).^67^ Comparing these estimates with the mentioned German study, we see again that the data by federal states correlate with the official German data. All these findings show that FB and Instagram undoubtedly capture valuable socio-demographic insights on Ukrainian refugees and that this data source is of great use in a situation where there is no official data. In the following (Figure 4), we tested the correlation between the number of Refugees from Ukraine in Poland and Facebook and Instagram users in Ukrainian in Poland.

**Figure 4.**
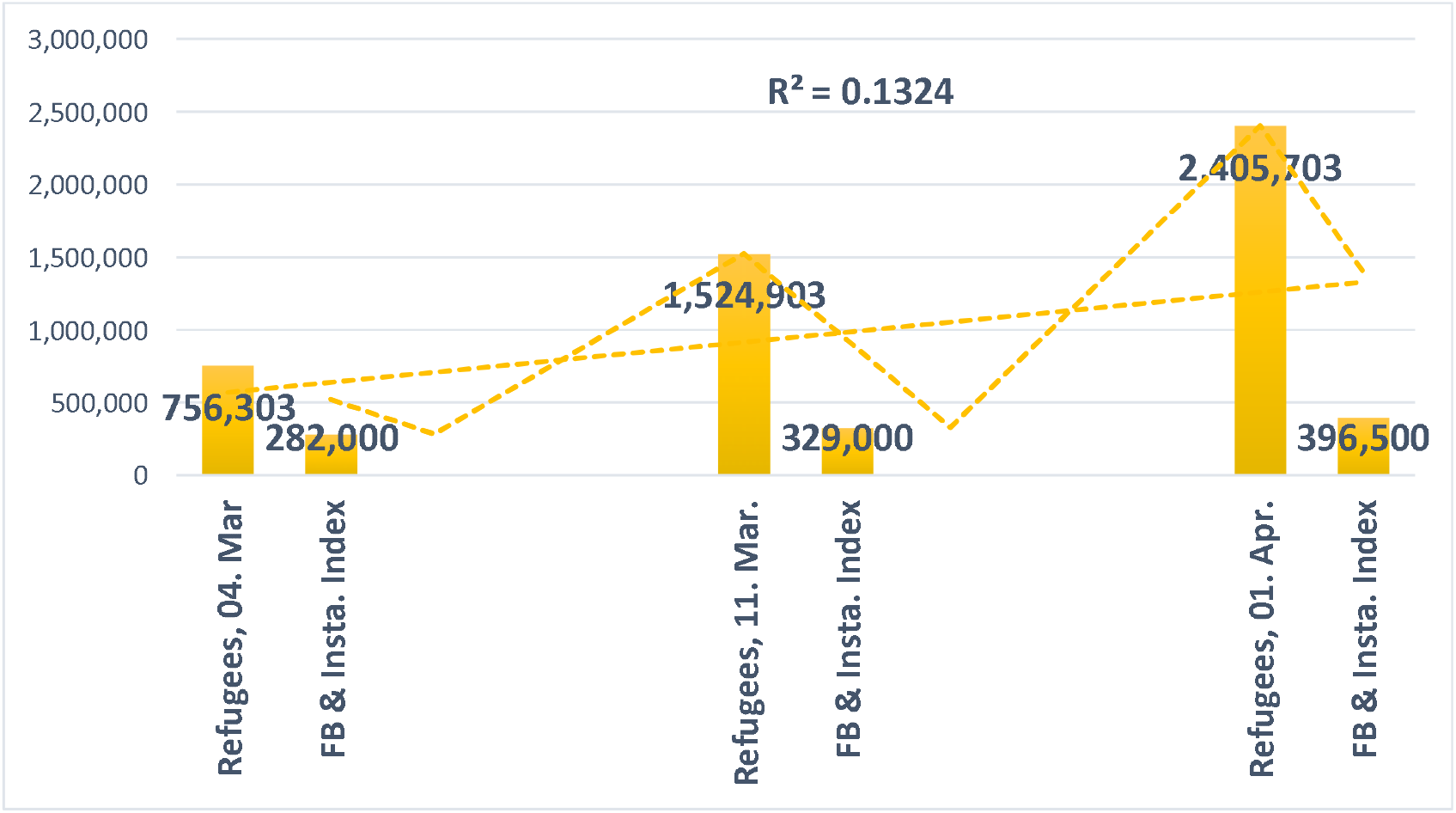
Correlation between Refugees from Ukraine in Poland and Facebook and Instagram users in Ukrainian in Poland on 04. and 11. March and 01. April 2022.

This tested correlation shows that the increase in the FB and Instagram index frequency is correlated with stepped-up emigration from Ukraine. R2 is 0.1324 and shows a positive correlation, and a p-value is statistically significant (Table 8).

**Table 8.**
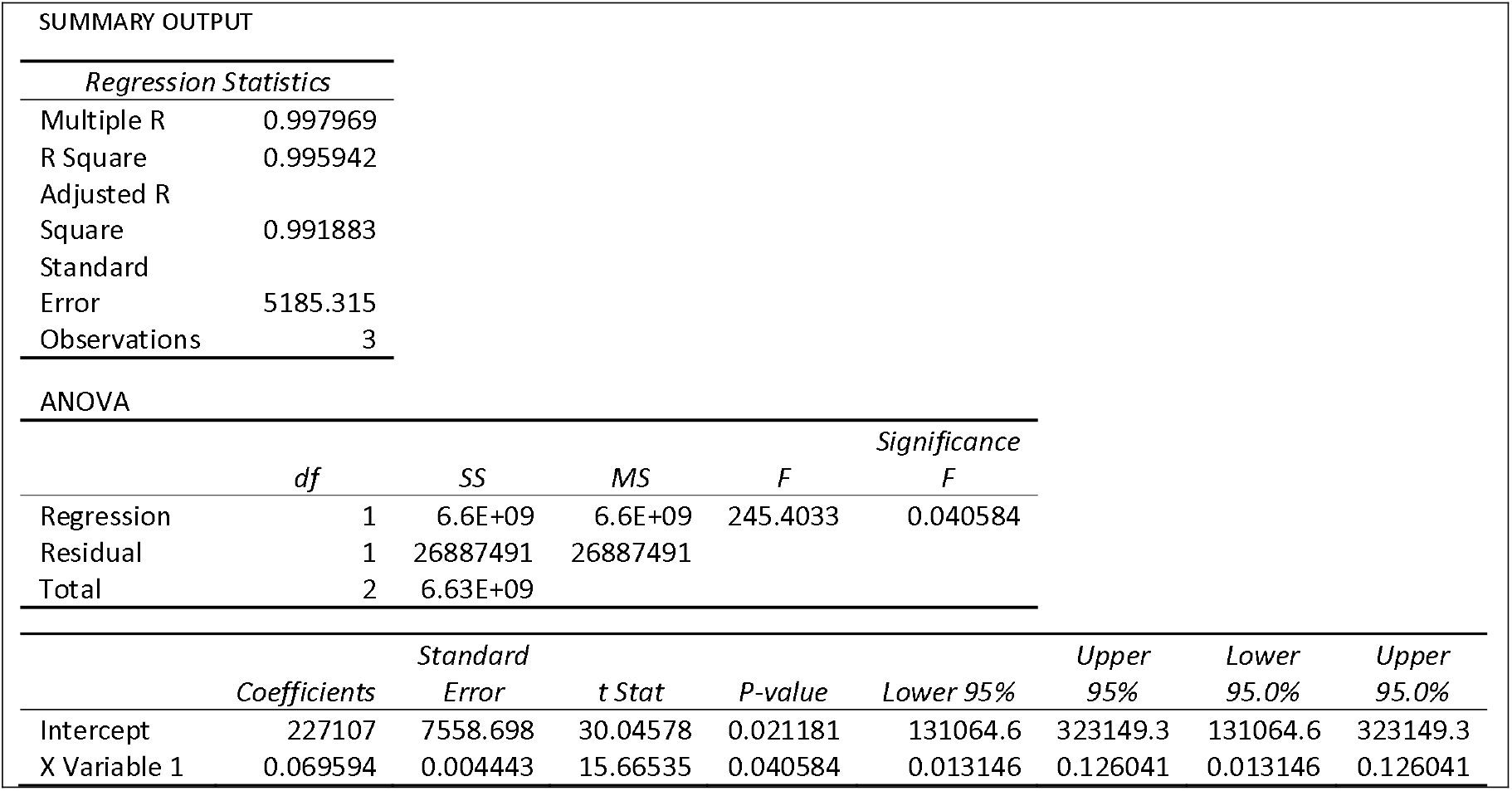
Regression analysis ANOVA.

**Table 9.**
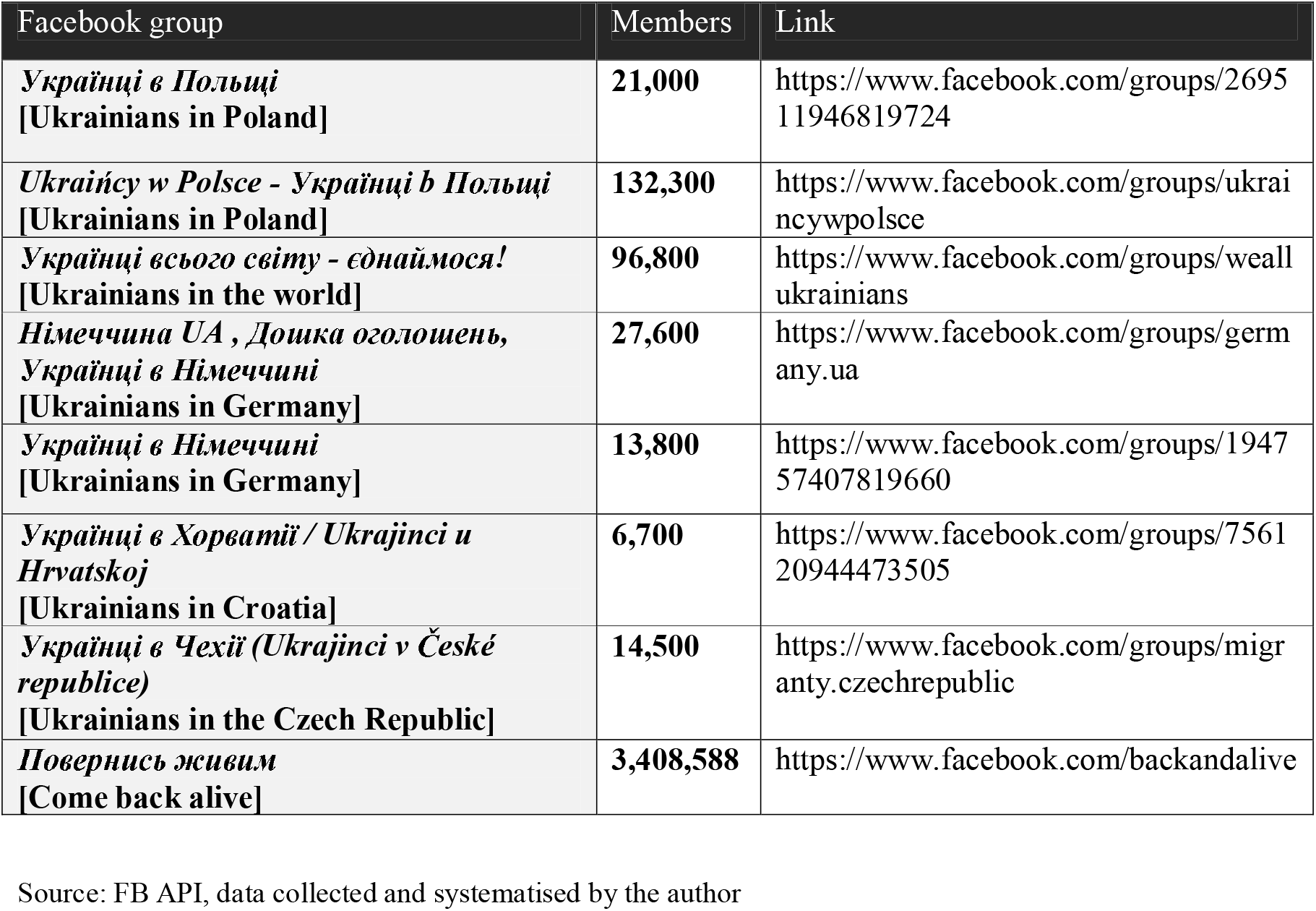
Facebook groups of Ukrainians in the EU as a source of socio-demographic data.

### V.2. Facebook and Instagram as a source of integration insights

Like many other diaspora groups, the Ukrainian diaspora, through new means of communication on the Internet, especially numerous FB groups, is creating a new form of sociability in the virtual space. In this way, a new form of social communities and collective identities is generated, the so-called “virtual diaspora identities”.^68^ In the virtual world of the diaspora community, FB groups become a place that unites and connects the diaspora and satisfies the need for social contact, which is often lacking in the first stages of emigrant life. The “imagining the homeland” plays a vital role in integrating into a new environment.^69^

Here we present some Ukrainian diaspora FB groups that we have analysed regarding the following questions: Is the number of new users growing? Are users looking for information about living abroad, etc.?

From analyses of the FB group members of the Ukrainian diaspora in Germany, Poland, Hungary, Czech Republic and Croatia, those groups serve as an essential source of information for those who plan to emigrate. An increase in the number of members from 1 March to 14 April was also noticed. These FB groups allow efficient exchange of information with compatriots who have already emigrated, which often facilitates the intention to emigrate to this specific country by many other refugees. In the following, we will analyse this approach in the example of Germany as a case study.

In Germany, numerous FB groups and portals are created where Ukrainian citizens exchange information about life in Germany in the Ukrainian language and maintain contact with compatriots from Ukraine. In our analysis of the FB group Українці в Німеччині (Ukrainians in Germany)^70^, we noticed that most of the comments and life experiences of this FB group confirm that moving to Germany was a good decision for many members, which is a strong message to those who plan emigration to Germany. Another way to use this data source to assess immigrant integration is to analyse users’ interests on their FB profiles. Table 6 shows the most frequently observed interests of Ukrainians citizens in Germany, according to FB.

The top interests of FB users in the Ukrainian language in Germany (Table 10) show that Ukrainian refugee interests differ from the interests of the German population. Nevertheless, it is useful to present this table here with current values so that we can see how much there has been a deviation to the interests of the German population in the future. For example, in the case of Croatian immigrants, the rapprochement of the interests of Croatians to the Germans was visible after three years.^71^ What is noticeable is that Ukrainians are increasingly expressing interest in German language learning sites, and this is undoubted a strong indicator of integration willingness of Ukrainian refugees into German society.

**Table 10.**
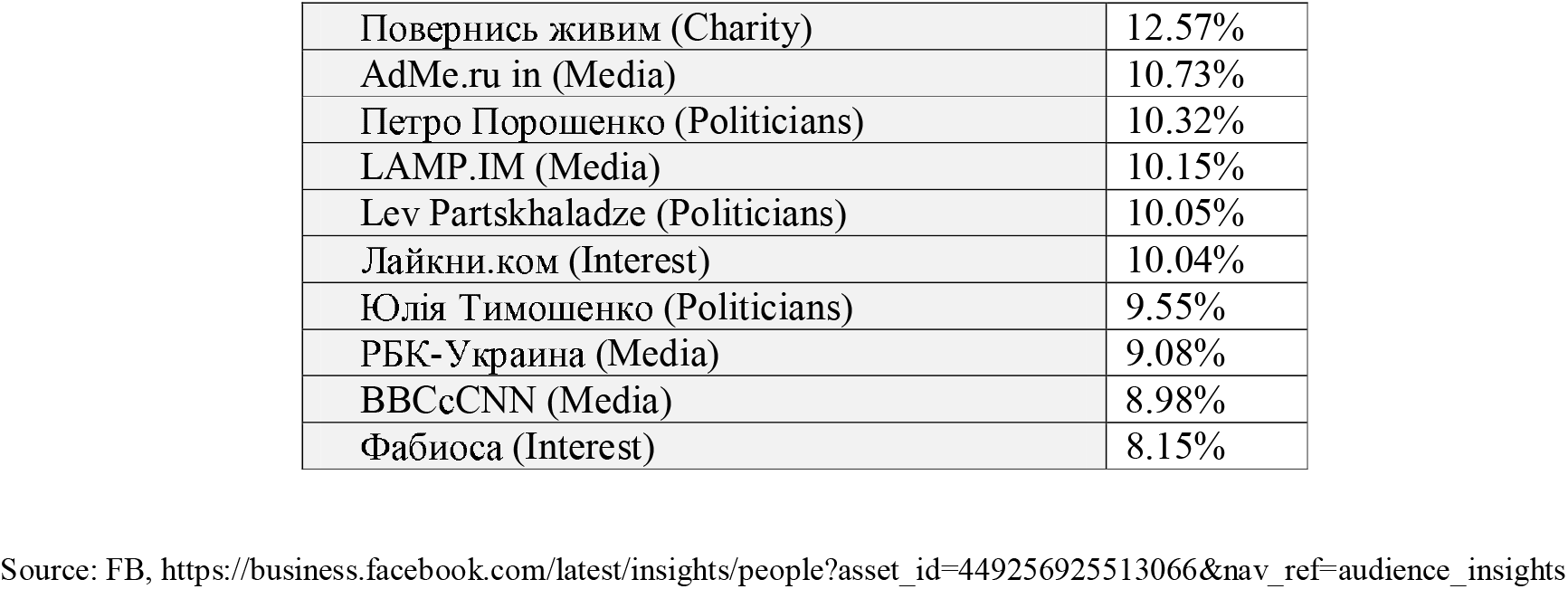
The most frequently observed interests of Ukrainians citizens in Germany, according to Facebook.

|  |  |
| --- | --- |
| Повернись живим (Charity) | 12.57% |
| AdMe.ru in (Media) | 10.73% |
| Петро Порошенко (Politicians) | 10.32% |
| LAMP.ІМ (Media) | 10.15% |
| Lev Partskhaladze (Politicians) | 10.05% |
| Лайкни.ком (Interest) | 10.04% |
| Юлія Тимошенко (Politicians) | 9.55% |
| РБК-Україна (Media) | 9.08% |
| ВВСсCNN (Media) | 8.98% |
| Фабиоса (Interest) | 8.15% |

Source: FB, https://business.facebook.com/latest/insights/people?asset_id=449256925513066&nav_ref=audience_insights

Jurić has shown that one can speak about high grad of integration willingness when regarding expressed interests one of the first three places is occupied by the pages where the German language can be learned. ^72^ As a simplified indication of the integration willingness of Ukrainians into German society in the future can serve interests typical for German society, such as German media, Web portals, series, films, music, etc.^73^ If the Ukrainians in Germany will have a similar level of interest as the Germans, this can be seen as an indicator of integration in terms of cultural taste.^74^ Since it is still too early to collect data of this type, in this first phase, the most reliable data on the integration willingness are available by monitoring the expression of interest of users of FB and Instagram in the Ukrainian language about Germany and learning German (Table 10).

Table 11 shows a rapid increase in interest in Germany and learning German by users of FB and Instagram in the Ukrainian language, especially in the period from 23. March to 03. April, when the increased movement of refugees from Ukraine by the UNHCR^75^ was registered. This observed increase in interest in Germany and learning German supports our thesis that many refugees intend to come to Germany.

**Table 11.** Interests of users of FB and Instagram in the Ukrainian language about Germany and learning German by countries of emigration (23. March -15. April 2022)

| **Year**<br>**2022** | **Poland** | **Hungary** | **Slovakia** | **Moldova** | **Romania** | **Germany** |
| --- | --- | --- | --- | --- | --- | --- |
| 23. Mar | *Deutsch*<br>*43,400*<br>*Deutschland*<br>*99,100* | *(no data*<br>*available) \** | *(no data*<br>*available) \** | *(no data*<br>*available) \** | *(no data*<br>*available) \** | *Deutsch*<br>*2,500*<br>*Deutschland*<br>*28,400* |
| 03. April | *Deutsch*<br>*67,000*<br>*Deutschland*<br>*154,400* | *Deutsch*<br>*2,200*<br>*Deutschland*<br>*6,000* | *Deutsch*<br>*2,700*<br>*Deutschland*<br>*7,400* | *Deutsch*<br>*6,700*<br>*Deutschland*<br>*8,800* | *Deutsch*<br>*(no data)*<br>*Deutschland*<br>*1,400* | *Deutsch*<br>*33,200*<br>*Deutschland*<br>*56,700* |
| 05. April | *Deutsch*<br>*67,100*<br>*Deutschland*<br>*155,600* | *Ibid* | *Ibid* | *Ibid* | *Ibid* | *Deutsch*<br>*33,300*<br>*Deutschland*<br>*56,800* |
| 15. April | *Deutsch*<br>*67,200*<br>*Deutschland*<br>*155,600* | *Deutsch*<br>*2,100*<br>*Deutschland*<br>*6,000* | *Ibid* | *Ibid.* | *Deutsch*<br>*under 1,000*<br>*(no data)*<br>*Deutschland*<br>*1,300* | *Ibid.* |

\* Note: Meta targeting does not show data unless it exceeds 1,000 users. Data collected and systematised by the author

### V.3. YouTube as a source of migration and integration data

In the Ukrainian refugee crisis, we start from an assumption that informing by watching videos on YouTube Platform from Ukraine about Poland or German cities, cultures and way of life gives reliable indications of the intention to migrate, i.e. flee to these countries. For this purpose, we analysed the search for interests related to life in Germany, videos about German cities, video lessons that offer the opportunity to learn German, etc. One of the contributions of this method is that it shows that by searching for video material on the YouTube platform, the intention of users to migrate, or in this case, to flee from Ukraine, can be estimated.

Figure 5 shows a high increase in interest in *Germany* and videos related to the so-called “German way of life”, which we interpreted as indicators of the intention of refugees to move to Germany.

**Figure 5.**
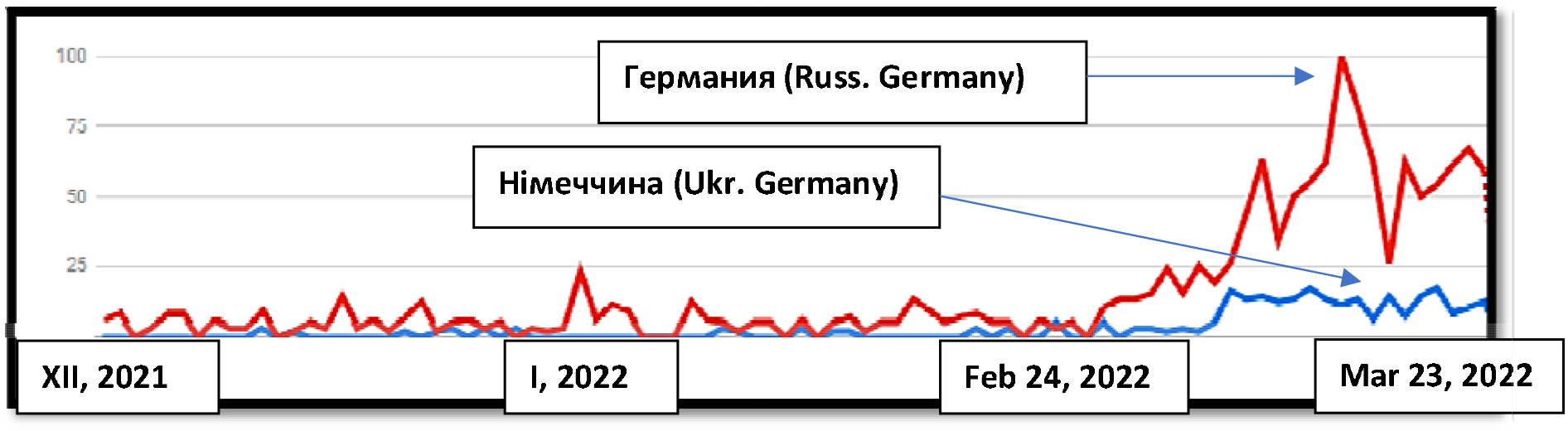
YouTube searches in Ukrainian and Russian related to life in Germany in Poland from 23. December 2021 to 23. March 2022.

**Figure 6.**
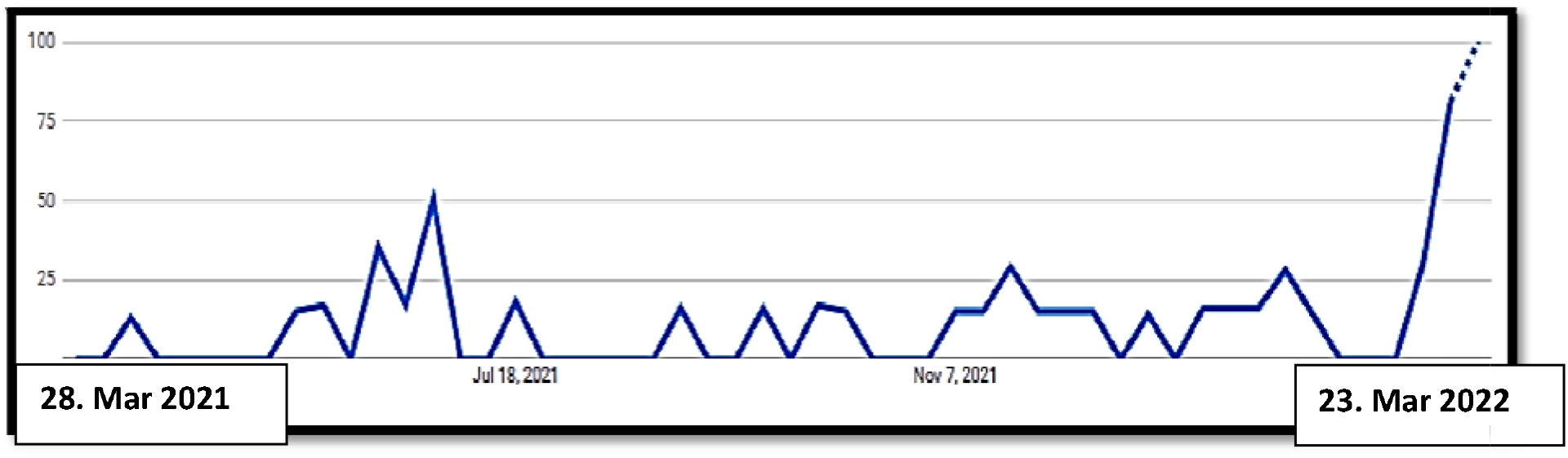
YouTube searches in Russian “Германия” (Germany) in Poland from 28. March 2021 to 23. March 2022.

As mentioned, Ukrainian refugee uses both Ukrainian and Russian during Internet searches. We have explained this phenomenon in the section Limitations. Above this explanation, it is possible that the citizens of Ukraine probably expect more information in Russian. Figure 7 shows that this trend is mainly observed after the war outbreak in Ukraine.

**Figure 7.**
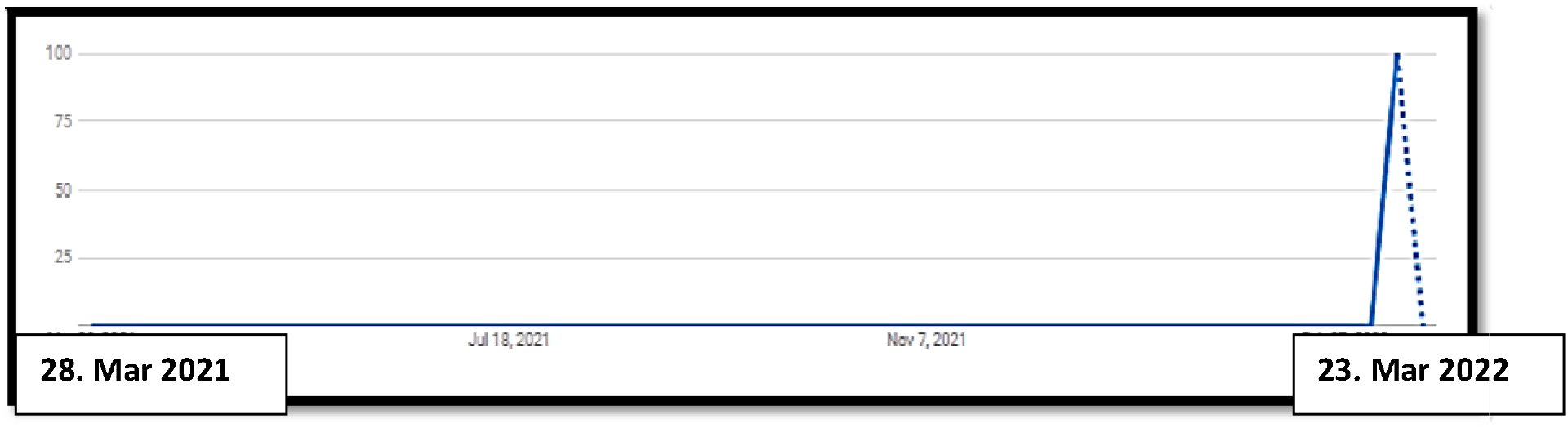
YouTube searches in Ukrainian “Німеччина” (Germany) in Slovakia from 28. March 2021 to 23. March 2022.

To test this method, we have applied it also to countries that, by all indications, are not the primary target of Ukrainian refugees, such as Slovakia and Croatia. Figure 8 shows that activity on YouTube in Ukrainian in Slovakia and Croatia has risen after the war outbreak in Ukraine.

**Figure 8.**
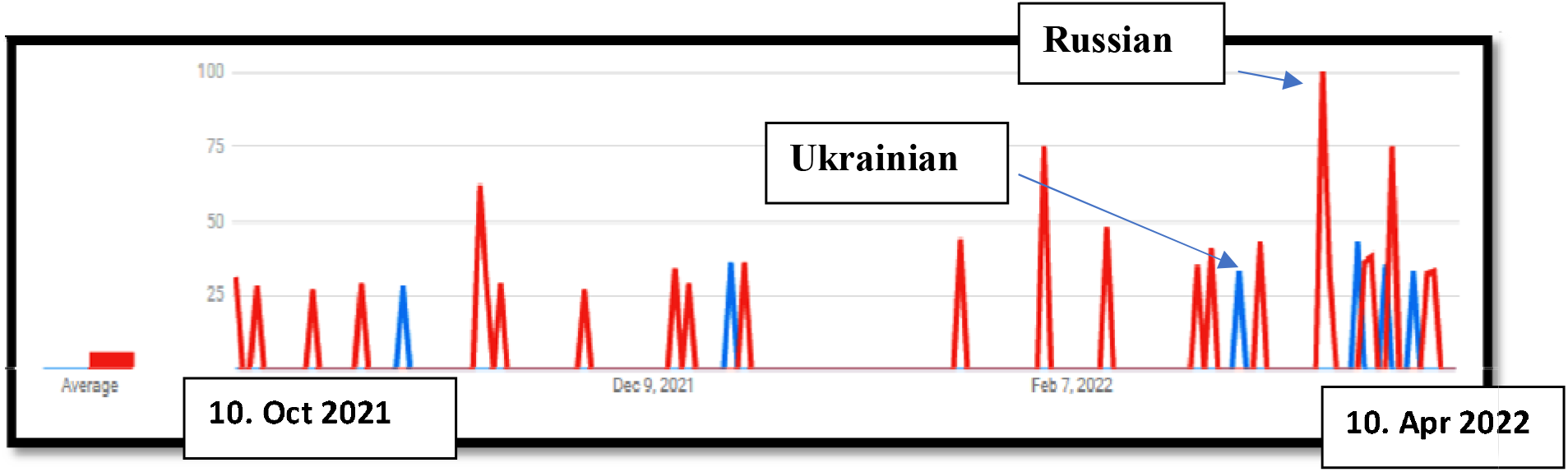
YouTube searches in Ukrainian “Хорватія” and Russian “Хорватия” (Croatia) in Croatia from 10. October 2021 to 10. April 2022.

In the case of Croatia, it can be noticed that the activity is primarily located in the coastal part of Croatia, mainly in Istria, Zadar and Split, which overlaps with Croatian media reports on the distribution of refugees in the country.

The data for Germany are shown below (Figures 9 and 10). Figure 9 shows a rapid increase in video searching trends in Russian about Germany after the outbreak of the war in Ukraine, which is a clear indicator of new users, i.e. refugees from Ukraine. Namely, no one else, or very few, would search in Russian or Ukrainian because it is logical that the requested materials are much more available in German.

**Figure 9.**
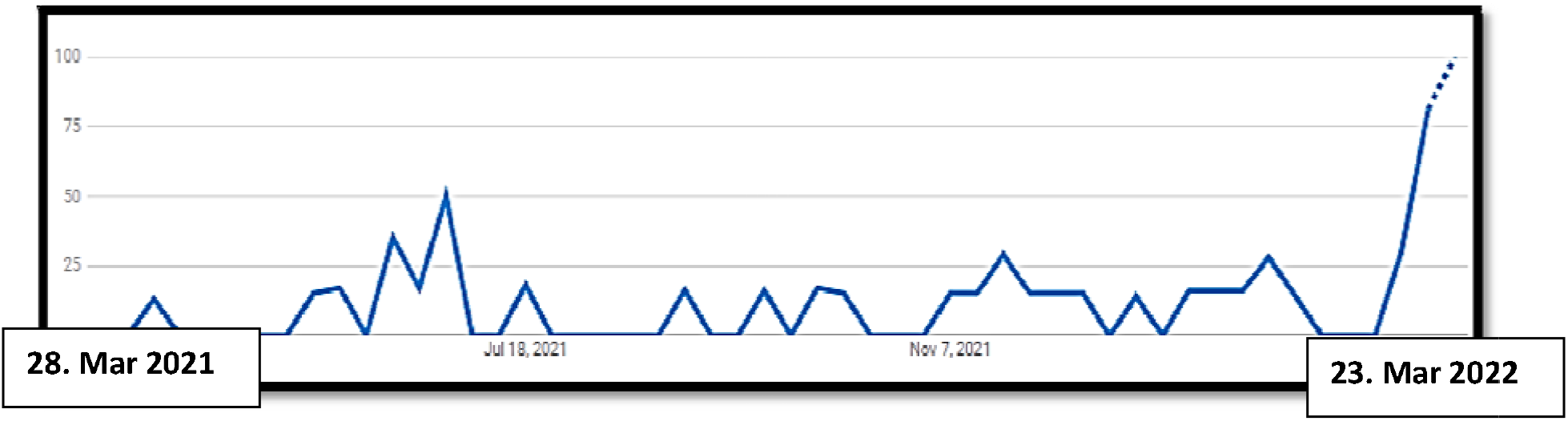
YouTube searches in Russian “Германия” (Germany) in Germany from 28. March 2021 to 23. March 2022.

**Figure 10.**
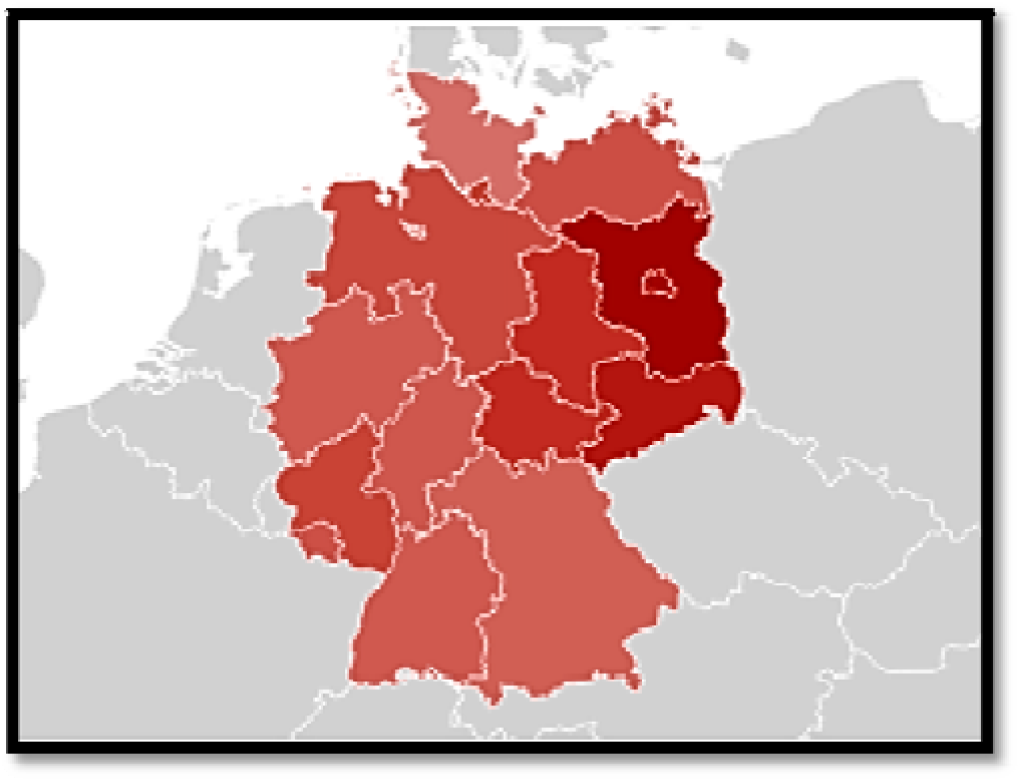
Overlap of YouTube data and official data on the prevalence of Ukrainian refugees in Germany. Note: darker red indicates a higher percentage of Ukrainian-language searches in Germany.

Figure 10 shows that the YouTube searches with the query “Germany” in Ukrainian and Russian correlate with the regions where the most Ukrainian refugees came in Germany.

YouTube can also provide several clues when it comes to integration. For example, Figure 11 shows an increased interest in learning German among Ukrainian refugees.

**Figure 11.**
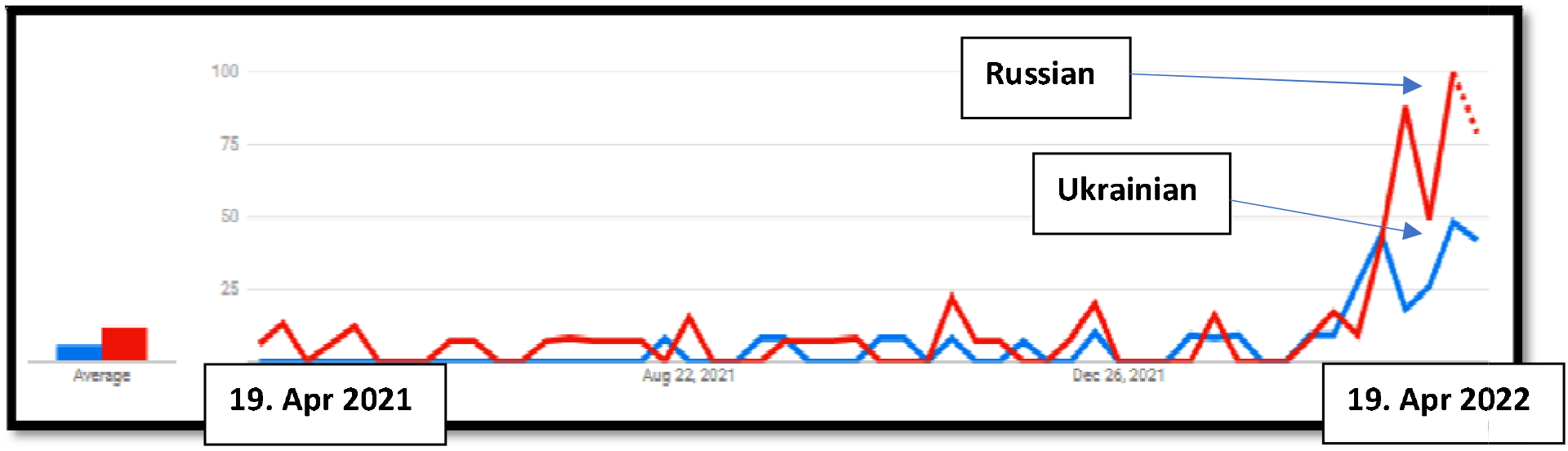
YouTube searches in Ukrainian “Вивчення німецької мови” and Russian “Изучение немецкого языка” (Learning German) in Germany from April 2021 to April 2022.

All the above examples indicate that the trend of emigration, i.e. fleeing from Ukraine, is still on the rise. At the very end, we mention one more useful social network – Twitter, which with its conversation tracking option, enables monitoring of the increase in interactions and the growth of interest in topics related to migration.^76^ Geo-referenced social media posts and micro-blogs, particularly geo-tagged Tweets, have been a popular source to track forced displacement streams.^77^ The basic idea is to use forced displaced people’s social media activities as a tracking device for their movements or new place of residence.^78^ A significant limitation is that only around 3% of all Tweets are geo-tagged.^79^ Tweets provide information on the location, nationality, birthplace, and the user’s language chosen when generating their account.

## VI. Limitations

Although previous research has shown the feasibility of using Big Data for migration studies,^80^ many open methodological issues exist.^81^ The primary limitations regarding FB and Instagram data in this study are as follows: Those data are not representative, they depend on Internet penetration rate in a specific group and do not include all age groups equally (for example, due to GDPR provisions, children are not included at all). The problem also lies in the possibility that users have multiple unlinked FB and Instagram accounts, leading to data distortion. However, after 2018, such practices have been significantly reduced after the scandal with Cambridge Analytica^82^ because FB then shut down millions of fake profiles^83^ and in suspicious cases, it requires verification of personal information.

Many FB users are our target group but are using FB, Instagram and YouTube in Russian. This phenomenon is numerous: During the 19^th^ century, the Russian government and in the 20^th^ century, the Soviet Union promoted the spread of the Russian language among the native Ukrainian population by suppressing the Ukrainian.^84^ In independent Ukraine, although Russian is not an official language, it is widely spoken, particularly in regions of Ukraine where Soviet Russification policies were the strongest, notably most of the urban areas of the east and south (Donetsk, Luhansk and Crimea).^85^

It is also necessary to consider the overall social context and that even before the war, many Ukrainians emigrated to Poland. 1,4 million Ukrainians lived in Poland before the war broke out (most fled after the Crimea crisis).^86^

Using the FB and the Instagram analytical tool is especially problematic because it provides data only for the current period, so it is necessary to monitor changes every day and keep your own data archive. By YouTube data, we do not know the exact number of searches, then just trends. From an ethical point of view, it is questionable how ethical it is to use data that was not collected with user consent^87^ (even when no personal data is known, as in this study).

On the other hand, the advantage of this approach is that it allows obtaining data on vulnerable groups for crisis management without further traumatising respondents who are inherent in traditional interview methods. Moreover, some of the presented issues, such as representativeness, are not unique to the presented method. Furthermore, new research confirms that the analysis of new forms of communication and social media reflects the existing social structures very well.^88^

It is encouraging that more and more researchers are perfecting this method^89^ and that both Google (YouTube) and Meta (FB and Instagram) realise specific data collection problems and are improving their methods of data collection. Combining traditional and new data sources is certainly key to making progress in migration and refugee studies.

## Conclusion

This study’s findings show that FB and Instagram undoubtedly capture valuable socio-demographic insights on Ukrainian refugees and that this data source is of great use in a situation where there is no official data and can serve crisis management. The usefulness and the main advantage of this approach are enabling first insights into integration willingness and identification of trends in the movement and intentions of refugees.

Testing performed matches the trend of immigration of Ukrainian refugees in Poland and Germany and confirms that FB and Instagram correctly notice trends. Regarding results of the cities and the German *Bundesländer* with the highest number of Ukrainian refugees in Germany, findings correspond to what official German statistics are reporting regarding socio-demographic data.

The tested correlation between the number of Ukrainian refugees in Poland and FB and Instagram users in Ukrainian in Poland shows that the increase in the frequency of the FB and Instagram index is correlated with stepped-up emigration from Ukraine. R2 is 0.1324 and shows a positive correlation, and a p-value is statistically significant.

The analysis of the FB group of Ukrainian diasporas in the EU shows that those FB groups can be a valuable source for studying integration. Ukrainians are increasingly expressing interest in German language learning sites, which is a strong indicator of the integration willingness of Ukrainian refugees into German society.

Regarding the second method we used, YouTube Insights tools, we showed that informing by watching videos on the YouTube platform from Ukraine about Poland or German cities, culture and way of life gives reliable indications of the intention to migrate, i.e. flee to these countries. Furthermore, the rapid increase in video searching trends in Ukrainian and Russian languages in Germany is a clear indicator of new users, i.e. refugees from Ukraine. This study also shows that the YouTube searches with the query “Germany” in Ukrainian and Russian correlate with the regions where the most Ukrainian refugees are in Germany. YouTube can also provide several clues when it comes to integration because video lessons that offer the opportunity to learn German give indices about the integration willingness of Ukrainian refugees in Germany. For example, YouTube searches in Ukrainian “Вивчення німецької мови” and Russian “Изучение немецкого языка” (Learning German) in Germany from April 2021 to April 2022 shows a quadruple increase in interest.

However, we are aware that this approach has many limitations and that many more studies are needed to perfect this method. Despite its limitations, we believe that the model presented here can contribute to migration and refuge studies.

## Compliance with ethical standards

In this work, we use only anonymous, aggregate data. All data are collected following the applicable GDPR and ethical principles of personal data handling.

## Data Availability

All data produced in the present work are contained in the manuscript

## Footnotes

1 UNHCR, Syria RRR, https://data2.unhcr.org/en/situations/Syria (08.03.2022)

2 Jurić, T. (2022). Predicting refugee flows from Ukraine with an approach to Big (Crisis) Data …

3 UNHCR Global Data Service (2021). Big (Crisis) Data for Predictive Models. A Literature Review, p 11.

4 Statista.com, https://www.statista.com/statistics/272014/global-social-networks-ranked-by-number-of-users/ (09.04.2022)

5 See: Jurić, T. (2022). Facebook i Google kao empirijska osnova za razvoj metode digitalnog praćenja vanjskih migracija hrvatskih građana, Ekonomski pregled, Vol. 73 No. 2, 2022., https://doi.org/10.32910/ep.73.2.2, https://hrcak.srce.hr/clanak/398317

6 Jurić, T. (2022). Forecasting Migration and Integration Trends Using Digital Demography – A Case Study of Emigration Flows from Croatia to Austria and Germany, Comparative Southeast European Studies 2022 https://doi.org/10.1515/soeu-2021-0090

7 Wladyka, D. K. 2017. “Queries to Google Search as Predictors of Migration Flows from Latin America to Spain.” Journal of Population and Social Studies 25 (4): 312–27.

8 Singh et al. (2019). KDD ‘19: Proceedings of the 25th ACM SIGKDD International Conference on Knowledge Discovery & Data Mining, p. 1975–1983, https://doi.org/10.1145/3292500.3330774

9 Connor, P. (2017). The digital footprint of Europe’s refugees. Methodology. Pew Research Center. 2017. URL: https://www.pewglobal.org/2017/06/08/online-searches-eu-refugees-methodology/ [20.03.2022]; Wanner P. (2020). How well can we estimate immigration trends using Google data?, Quality & Quantity, 2020. https://doi.org/10.1007/s11135-020-01047-w

10 Ibáñez Sales, M. (2021). Big data at the crossroads: seizing the potential of Big data to guide the future of EU migration policy. Euromesco Policy brief n. 116, p. 2.

11 UNHCR Global Data Service (2021). Big (Crisis) Data for Predictive Models. A Literature Review, p 11.

12 Jurić, T. (2022). Forecasting Migration and Integration Trends Using Digital Demography …

13 Zagheni, E., M. Polimis, M. Alexander, I. Weber, and F. C. Billari. 2020. “Combining Social Media Data and Traditional Surveys to Nowcast Migration Stocks in the United States.” Population Research and Policy Review, https://doi.org/10.1007/s11113-020-09599-3.

14 Zagheni, E., I. Weber, and K. Gummadi. 2017. “Leveraging Facebook’s Advertising Platform to Monitor Stocks of Migrants.” Population and Development Review 43 (4): 721–34.

15 Cesare, N., H. Lee, T. McCormick, E. Spiro, and E. Zagheni. 2018. “Promises and Pitfalls of UsinDigital Traces for Demographic Research.” Demography 55: 1979–99.; Schneider, D., and K. Harknett. 2019. “What’s to Like? Facebook as a Tool for Survey Data Collection.” Sociological Methods & Research 20 (10): 1–33.

16 Jurić, T. (2022). Predicting refugee flows from Ukraine with an approach to Big (Crisis) Data …

17 See: Jurić, T. (2022). Forecasting Migration and Integration Trends Using Digital Demography …

18 Dubois, A., E. Zagheni, K. Garimella, and I. Weber. 2018. “Studying Migrant Assimilation Through Facebook Interests.” Lecture Notes in Computer Science 11186: 51–60.

19 Herdagdelen, A., B. State, L. Adamic, and W. Mason. 2016. “The Social Ties of Immigrant Communities in the United States.” In WebSci 16: Proceedings of the 8th ACM Conference on Web Science, 78–84. New York: Association for Computing Machinery.

20 Jurić, T. (2022). Forecasting Migration and Integration Trends Using Digital Demography …

21 Meta Business Suite, https://business.facebook.com/latest/insights/people?asset_id=449256925513066&nav_ref=audience_insights

22 Meta Business Suite, ibid.

23 Jurić, T. (2022). Facebook i Google kao empirijska osnova za razvoj metode digitalnog praćenja vanjskih migracija hrvatskih građana, https://doi.org/10.32910/ep.73.2.2,

24 Ibid.

25 Jurić, T. (2022). Forecasting Migration and Integration Trends Using Digital Demography …

26 Wanner P. (2020). How well can we estimate immigration trends using Google data?, Quality & Quantity, 2020. https://doi.org/10.1007/s11135-020-01047-w

27 Jurić, T. (2022). Predicting refugee flows from Ukraine with an approach to Big (Crisis) Data: a new opportunity for refugee and humanitarian studies, doi: https://doi.org/10.1101/2022.03.15.22272428.

28 See: Böhme, M. B., A. Gröger, and T. Stöhr. 2020. “Searching for a Better Life: Predicting International Migration with Online Search Keywords.” Journal of Development Economics 142, https://doi.org/10.1016/j.jdeveco.2019.04.002.; Wanner, P. 2020. “How Well CanWeEstimate Immigration Trends Using Google Data?” Quality and Quantity 55: 1181–202.; Wladyka, D. K. 2017. “Queries to Google Search as Predictors of Migration Flows from Latin America to Spain.” Journal of Population and Social Studies 25 (4): 312–27.

29 Jurić, T. (2022). Forecasting Migration and Integration Trends Using Digital Demography …

30 Alba, Richard; Nee, Victor (1997). “Rethinking Assimilation Theory for a New Era of Immigration”. International Migration Review. 31, 4 (4): 826–874. doi:10.1177/019791839703100403.

31 UNHCR, The Integration of Refugees. A Discussion Paper (2018). https://www.unhcr.org uploads› sites› 2018/02 (16.04.2022)

32 Strang, A., and A. Ager. “Refugee Integration: Emerging Trends and Remaining Agendas.” Journal of Refugee Studies 23, no. 4 (2010): 589–607. doi:10.1093/JRS/FEQ046.

33 UNHCR, The Integration of Refugees. A Discussion Paper (2018). https://www.unhcr.org uploads› sites› 2018/02 (16.04.2022)

34 Ibid.

35 See: Jurić, T. (2022). Forecasting Migration and Integration Trends Using Digital Demography – A Case Study of Emigration Flows from Croatia to Austria and Germany, https://doi.org/10.1515/soeu-2021-0090

36 Jurić, T. (2022). Forecasting Migration and Integration Trends Using Digital Demography …

37 Statcounter, https://gs.statcounter.com/social-media-stats/all/Ukraine (09.04.2022)

38 Statista.com, https://www.statista.com/statistics/1030052/facebook-users-ukraine/ (09.04.2022)

39 Ibid.

40 Wanner, (Böhme, Gröger, and Stöhr 2020; Wilde, Chen, and Lohmann 2020)

41 Jurić, T. (2021). *“Gastarbeiter Millennials”. Exploring the past, present and future of migration from Southeast Europe to Germany and Austria with approaches to classical, historical and digital demography*. Hamburg 2021, Verlag Dr. Kovač. p 260.

42 Zagheni, E., I. Weber, K.Gummadi, (2017). Leveraging Facebook’s advertising platform to monitor stocks of migrants, Population and development review 0(0): 1–14 (xxx 2017)

43 FB: https://developers.facebook.com/docs/marketing-api/buying-api/targeting (accessed 16.07.2020)

44 Dubois, A., Zagheni E, Garimella, I. Weber, (2018). Studying Migrant Assimilation Through Facebook Interests, Springer

45 Facebook, Geschätzte Zielgruppengröße, https://www.facebook.com/business/help/1665333080167380?id=176276233019487 (09.04.2022)

46 UNHCR, Ukraine Situation Regional Refugee Response Plan, https://data2.unhcr.org/en/situations/Ukraine (04.03.2022)

47 UNHCR, Syria RRR, https://data2.unhcr.org/en/situations/Syria (08.03.2022)

48 Jurić, T. (2022). Predicting refugee flows from Ukraine with an approach to Big (Crisis) Data …

49 IOM, Situation Report Ukraine, https://www.iom.int/news/almost-65-million-people-internally-displaced-ukraine-iom

50 UNICEF, https://news.un.org/en/story/2022/03/1114592

51 IOM, Situation Report Ukraine.

52 European Commission, Ukraine: Commission proposes temporary protection for people fleeing war in Ukraine and guidelines for border checks, https://ec.europa.eu/commission/presscorner/detail/en/ip_22_1469

53 UNHCR, https://data2.unhcr.org/en/situations/ukraine

54 Medendienst Integration, https://mediendienst-integration.de/migration/flucht-asyl/ukrainischefluechtlinge.html (01.04.2022)

55 UNHCR, https://data2.unhcr.org/en/situations/ukraine (08.03.2022)

56 Bundesministerium des Innern und für Heimat, Pressemitteilung, 04.04.2022, Befragung ukrainischer Kriegsflüchtlinge, https://www.bmi.bund.de/SharedDocs/downloads/DE/veroeffentlichungen/nachrichten/2022/umfrage-ukraine-fluechtlinge.pdf?blob=publicationFile&v=2

57 Jurić, T. (2022). Predicting refugee flows from Ukraine with an approach to Big (Crisis) Data …

58 Medendienst Integration, https://mediendienst-integration.de/migration/flucht-asyl/ukrainische-fluechtlinge.html (10.04.2022)

59 Bundesministerium des Innern und für Heimat, Pressemitteilung, 04.04.2022, Befragung ukrainischer Kriegsflüchtlinge.

60 Medendienst Integration, https://mediendienst-integration.de/migration/flucht-asyl/ukrainische-fluechtlinge.html (10.04.2022)

61 Medendienst Integration, https://mediendienst-integration.de/migration/flucht-asyl/ukrainische-fluechtlinge.html (10.04.2022)

62 Bundesministerium des Innern und für Heimat, Pressemitteilung, 04.04.2022, Befragung ukrainischer Kriegsflüchtlinge.

63 Germany: Grieshaber, Kirsten (17 March 2022). “Berlin train station turns into refugee town for Ukrainians”. Associated Press. Archived from the original on 18 March 2022. Retrieved 26 March 2022; Moldova: https://www.middleeasteye.net/news/russia-ukraine-war-moldova-greets-refugees-compassion-wariness; Poland: Gov.pl, Punkty recepcyjne - Urząd do Spraw Cudzoziemców - Portal Gov.pl”. Urząd do Spraw Cudzoziemców (in Polish), https://www.gov.pl/ (02.04.2022); Slovakia: Update: Initial Assessment Report: Eastern Slovakia 5th March 2022, https://reliefweb.int/report/slovakia/update-initial-assessment-report-eastern-slovakia-5th-march-2022; Romania: https://adevarul.ro/news/politica/aurescu-cnn-460000-refugiati-ucraineni-tranzitat-romania-80000-ramas-tara-noastra-1_6230e9bc5163ec427143ad8e/index.html

64 Bundesministerium des Innern und für Heimat, Pressemitteilung, 04.04.2022, Befragung ukrainischer Kriegsflüchtlinge, https://www.bmi.bund.de/SharedDocs/downloads/DE/veroeffentlichungen/nachrichten/2022/umfrage-ukraine-fluechtlinge.pdf?blob=publicationFile&v=2

65 Bundesministerium des Innern und für Heimat, Pressemitteilung, 04.04.2022, Befragung ukrainischer Kriegsflüchtlinge.

66 Bundesministerium des Innern und für Heimat, Pressemitteilung, 04.04.2022, Befragung ukrainischer Kriegsflüchtlinge.

67 Ibid.

68 See: Jurić, T. (2021). *“Gastarbeiter Millennials”. Exploring the past, present and future of migration from Southeast Europe to Germany and Austria with approaches to classical, historical and digital demography*. Hamburg 2021, Verlag Dr. Kovač. p 455.

69 Chan, B. (2005). Imagining the Homeland: The Internet and Diasporic Discourse of Nationalism, Journal of Communication Inquiry, https://doi.org/10.1177/0196859905278499

70 FB group Українці в Німеччині (Ukrainians in Germany), https://www.facebook.com/groups/germany.ua

71 Jurić, T. (2022). Forecasting Migration and Integration Trends Using Digital Demography …

72 Ibid.

73 See: D. Schneider and K. Harknett, What’s to like? Facebook as a tool for survey data collection,” Sociological Methods & Research, 2019.

74 See: Dubois et al., 2018.

75 UNHCR, https://data2.unhcr.org/en/situations/ukraine

76 Zagheni E., Garimella V.R.K., Weber I. and State B. (2014). Inferring International and Internal Migration Patterns from Twitter Data. 439-444.

77 UNHCR Global Data Service (2021). Big (Crisis) Data for Predictive Models. A Literature Review, p 19

78 Alina Sirbu, Gennady Andrienko, Natalia Andrienko, Chiara Boldrini, Marco Conti, Fosca Giannotti, Riccardo Guidotti, Simone Bertoli, Jisuand Kim, Cristina Ioana Muntean, Luca Pappalardo, Andrea Passarella, Dino Pedreschi, Laura Pollacci, FrancescaPratesi, and Rajesh Sharma. Human migration: the big data perspective. International Journal of Data Science and Analytics, March 2020.

79 Alessandra Righi. Assessing migration through social media: a review. Mathematical Population Studies, 2019.

80 Zagheni, E., Polimis, M. Alexander, I. Weber, F. C. Billari (2018). Combining Social Media Data and Traditional Surveys to Nowcast Migration Stocks.

81 See: Cesare et al., Promises and Pitfalls of Using Digital Traces for Demographic Research. Demography volume 55, pages1979–1999 (2018).

82 See: Vox, https://www.vox.com/policy-and-politics/2018/3/23/17151916/facebook-cambridge-analytica-trump-diagram (20.03.2022)

83 Jurić, T. (2022). Facebook i Google kao empirijska osnova za razvoj metode digitalnog praćenja vanjskih migracija hrvatskih građana, https://doi.org/10.32910/ep.73.2.2, https://hrcak.srce.hr/clanak/398317

84 Magoscy, R. (1996). A History of Ukraine. Toronto: University of Toronto Press.

85 Ibid.

86 The Guardian (2019), https://www.theguardian.com/world/2019/apr/18/whole-generation-has-gone-ukrainian-seek-better-life-poland-elect-president (14.04.2022)

87 Ibid.

88 Kalimeri, K., A. Bonanomi, M. Beiro, A. Rosina, and C. Cattuto. 2020. “Traditional versus Facebook-Based Surveys: Evaluation of Biases in Self-Reported Demographic and Psychometric Information.” Demographic Research 42: 133–48.

89 B. Zhang, M. Mildenberger, P. D. Howe, J. Marlon, S. A. Rosenthal, and A. Leiserowitz, Quota sampling using Facebook advertisements,” Political Science Research and Methods, pp. 1-7, 2018.; S. Pöotzschke and M. Braun, Migrant sampling using FB advertisements: A case study of Polish migrants in four European countries,” Social Science Computer Review, vol. 35, no. 5, pp. 633{653, 2017.

